# Early Detection of Emerging SARS-CoV-2 Variants of Interest for Experimental Evaluation

**DOI:** 10.1101/2022.08.08.22278553

**Authors:** Zachary S. Wallace, James Davis, Anna Maria Niewiadomska, Robert D. Olson, Maulik Shukla, Rick Stevens, Yun Zhang, Christian M. Zmasek, Richard H. Scheuermann

## Abstract

Since the beginning of the COVID-19 pandemic, SARS-CoV-2 has demonstrated its ability to rapidly and continuously evolve, leading to the emergence of thousands of different sequence variants, many with distinctive phenotypic properties. Fortunately, the broad availability of next generation sequencing (NGS) technologies across the globe has produced a wealth of SARS-CoV-2 genome sequences, offering a comprehensive picture of how this virus is evolving so that accurate diagnostics and reliable therapeutics for COVID-19 can be maintained. The millions of SARS-CoV-2 sequences deposited into genomic sequencing databases, including GenBank, BV-BRC, and GISAID are annotated with the dates and geographical regions of sample collection, and can be aligned to the Wuhan-Hu-1 reference genome to extract the constellation of nucleotide and amino acid substitutions. By aggregating these data into concise datasets, the spread of variants through space and time can be assessed. Variant tracking efforts have focused on the spike protein due to its critical role in viral tropism and antibody neutralization. To identify emerging variants of concern as early as possible, we developed a computational pipeline to process the genomic data from public databases and assign risk scores based on both epidemiological and functional parameters. Epidemiological dynamics are used to identify variants exhibiting substantial growth over time and across geographical regions. In addition, experimental data that quantify Spike protein regions critical for adaptive immunity are used to predict variants with consequential immunogenic or pathogenic impacts. These growth assessment and functional impact scores are combined to produce a *Composite Score* for any set of Spike substitutions detected. With this systematic approach to routinely score and rank emerging variants, we have established a method to identify threatening variants early and prioritize them for experimental evaluation.

## Introduction

The ongoing evolution of SARS-CoV-2 has remained a persistent public heath challenge throughout the entire course of the pandemic. In just two and a half years since the first strain of the virus was isolated and fully sequenced, SARS-CoV-2 has evolved into hundreds of thousands of strains containing unique combinations of mutations, also known as variant constellations, with many of these mutations leading to altered virus phenotypes in terms of antigenicity, transmissibility, and viral fitness.^25,26^ In order to label the rapidly growing collection of variants, the scientific community has relied on the PANGO Lineage^21^ nomenclature or WHO classifications as naming schemes for these variants. The continual emergence of SARS-CoV-2 variants has caused public health agencies to categorize these lineages depending on their predicted importance. In the United States, the CDC has identified these as four categories: *Variants Being Monitored (VBM), Variants of Interest (VOI), Variants of Concern (VOC),* or *Variants of High Consequence (VOHC)*. Classification is dependent on viral growth dynamics and level of threat to preexisting immunity or therapeutic efficacy. Notable *Variants of Concern* include B.1.1.7 (WHO class Alpha), B.1.617.2 (WHO class Delta), and the most recent B.1.1.529/BA.1-BA.5 (WHO class Omicron).^27^

It has been a consistent pattern throughout the course of the pandemic for SARS-CoV-2 to acquire genetic changes of functional and epidemiological importance, beginning with the observation as early as Spring 2020 that the Spike protein mutation, D614G, was associated with higher viral loads and was under positive selection.^16^ Since then, with the emergence of Alpha and Delta, Spike mutations such as N439R, N501Y, E484K, and P681H, have been linked to increased ACE2 binding affinity, antibody binding escape, and enhanced viral replication.^3,29,32^, The most recently emerged *Variant of Concern*, Omicron, has been reported to contain constellations of mutations across the n-terminal domain (NTD) and receptor binding domain (RBD). This has resulted in high levels of viral antigenic escape, and partial or complete resistance to the majority of available therapeutic monoclonal antibodies and, to a large extent, infection or vaccine-elicited polyclonal sera.^14,30^ The astonishing ability of SARS-CoV-2 to rapidly evolve into variants with expanded cellular tropism, enhanced replication, increased transmissibility, and evasion of preexisting immunity has triggered the scientific community to band together and critically monitor the evolution of this virus through efforts like the US National Institute of Allergy and Infectious Diseases (NIAID)’s SARS-CoV-2 Assessment of Viral Evolution (SAVE) program^1^ and COVID-19 Genomics UK Consortium (COG UK), which seek to iteratively provide a risk-assessment of emerging variants of interest and offer recommendations towards an optimal public health response.

A key component to successfully monitoring viral evolution, and thereby setting the stage for risk assessment, is through genomic surveillance.^12,16^ Thanks to the growing availability of next generation sequencing (NGS) technologies, SARS-CoV-2 genome sequences from millions of infected individuals across the globe have been determined and deposited into public databases^11^ such as the National Center for Biotechnology Innovation (NCBI) GenBank, the Global Initiative on Sharing Avian Influenza Data (GISAID)^20^, and the Bacterial and Viral Bioinformatics Resource Center (BV-BRC)^24^. These data have been dynamically growing at a rapid pace since the early stages of the pandemic and have allowed researchers to pinpoint mutations under positive or negative selection through the evaluation of occurrence rates of synonymous versus non-synonymous nucleotide substitutions or identify the mutational drivers of evolution by modeling growth as a linear combination of the effects of individual mutations.^15,31^ Overall, the amount of SARS-CoV-2 genomic sequencing being carried out on a global scale and the bioinformatics capabilities to understand these data has opened opportunities for diverse research into strategies for the *early detection of emerging variants of concern*. This is a concept by which computational frameworks are used to manage and analyze rapidly growing viral sequencing data and ultimately prioritize emerging variants based on epidemiological dynamics and functional characteristics in near real time^1,15,17^. The concept of *early detection* is a key aspect of the NIAID SAVE program - to select and prioritize the variants for *in vitro* and *in vivo* experimentation to assess risk of novel emerging variants (see Reference #1 for a complete description of the NIAID SAVE program for more details of the integrated workflow).

Here we present a computational heuristic developed for early detection of SARS-CoV-2 emerging variants of interest, which combines spatiotemporal sequence prevalence dynamics and geographic spread with functional impact prediction, that can be used to rank variants composed of constellations of substitutions. The methods described have been used to rank emerging variants of interest for the NIAID SAVE consortium Early Detection group every month since early 2021 to inform wet-lab experimentation.

## Methodology and Results

### Algorithms for Early Detection

#### Sequence Prevalence Dynamics

The BV-BRC team has developed the *SARS-CoV-2 Early Detection and Analysis Pipeline* to offer informatics support for analyzing emerging SARS-CoV- 2 variants from genomic sequencing data and associated epidemiological metadata processed from public databases. Throughout the pandemic, the team has run this pipeline nearly every week to process SARS-CoV-2 genomic sequencing and epidemiological data and support the Early Detection component of the NIAID SAVE program. Data for this pipeline can be downloaded from the EpiCov portal of GISAID^20^, NCBI GenBank, Virus Pathogen Resource^23^, or BV-BRC^24^ databases. A primary aspect of this pipeline is the ability to capture four sequence prevalence dynamics segregated by month and geographic location: 1) the total SARS-CoV-2 genome sequence counts, 2) the genome sequence counts for specific lineages and variants, 3) the sequence prevalence of these lineages and variants, and 4) the growth rates of these lineages and variants from month to month (see **Supplementary Material** for more details). These sequence prevalence dynamics are calculated for individual PANGO lineages^21^, single amino acid substitutions found in any SARS-CoV-2 protein, and unique constellations of protein substitutions denoted as “covariates”, which we refer to collectively as “variants”. Note that covariates consist of a string of single amino acid mutations identified in one or more proteins of some SARS-CoV-2 sequence isolate. Each covariate also belongs to a specific PANGO lineage based on the nomenclature status of the origin sequence. Multiple covariates of a single protein can belong to the same PANGO lineage. In this work, we focus on the analysis of Spike covariates since this has been the initial focus of the NIAID SAVE program.

Generating the sequence prevalence dynamics of emerging variants provides the required epidemiological parameters needed by the scoring heuristics for predicting which variants might be of concern. We designed a workflow to capture these dynamics as an upstream component of the scoring heuristics.

#### Capturing the Dynamics of Emerging Variants

A custom built pipeline was routinely used to compute sequence prevalence dynamics by capturing SARS-CoV-2 variants and isolation metadata (geographic location and date of isolation) from the genomic sequencing databases. Strict quality control criteria were used to filter out genomes with high numbers of ambiguous or indeterminate bases (n>x), low sequence length coverage (n<x,000kb), missing viral names, or improper metadata such as incorrect representation of dates or region names. Genomes were then pairwise aligned to the Wuhan-Hu-1 reference genome (NC_045512.2), and a constellation of variants was extracted for the Spike protein, or for the entire SARS-CoV-2 proteome (all 16 non-structural proteins as well as E, M, N, S, ORF3a, ORF6, ORF7a, ORF7b, ORF8, ORF9b, ORF10, and ORF14). Variant constellations can then be partitioned into temporal and geographic groups. The total number of genomes isolated for these groups is also calculated and serves as the denominator for the prevalence ratio (formula 1). Dates were then partitioned by month, and variant constellation counts and total isolate counts per region per date month were used to compute epidemiological dynamics, namely the prevalence and growth rates of variants by month as shown in formulas (1) and (2).

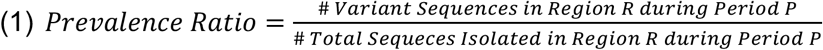

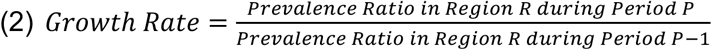

Running this process to identify variant constellations and acquire region-date counts directly from the raw sequence data is a very computationally expensive task for a large batch of genomes and requires High-Performance Computing. An alternative approach is to use preprocessed data provided in GISAID metadata files, which can be downloaded with a registered GISAID account. Records include variant constellations, geographic location, date of collection, and basic quality control data such as the sequence length and ambiguous nucleotide content. These files can be used to extract the Spike variant constellation of each record, count the occurrence of these Spike variants by region and date, bin the dates by month, and compute the monthly prevalence and growth rate dynamics for each Spike variant based on the total sequence record counts per region and date. PANGO lineage designations can also be used to count the occurrence of specific lineages by region and date and compute spatiotemporal dynamics. A similar strategy can be used to collect data for single amino acid substitutions rather than variant constellations or PANGO Lineages, iterating through each sequence record, and counting the occurrence of each single mutation by region and date and then computing the prevalence and growth rates of these single mutations by month per region.

This workflow was used to generate sequence prevalence dynamics based on either GISAID metadata files or GenBank/BV-BRC FASTA files and then fed into the scoring heuristics component of the pipeline (see the **Supplementary Material** for more details).

#### Sequence Prevalence Score

To identify SARS-CoV-2 variations with concerning epidemiological dynamics, a scoring heuristic based on sequence prevalence ratio (Formula 1), growth in sequence prevalence (Formula 2), and geographic spread was devised - the *Sequence Prevalence Score*. This scoring method can be applied to rank variants within the entire dataset, within a specific PANGO lineage, a specific WHO clade, or a user supplied list of covariates. First, this method segregates variants for each country. Next, only variants with a sequence count greater than 10 in the most recent month are retained to control for large apparent growth rates associated with small numbers. Any variants that were filtered out are assigned a score of 0. To calculate the Sequence Prevalence Score, the most recent three months of data are used. A score of 1 is assigned for every country/month combination in which the sequence prevalence is >5% or the growth rate from the previous month is >5-fold. These numbers are then summed to obtain the final Sequence Prevalence Score across all countries/month combinations. Three months of data is the default interval applied, but users can specify this parameter through a “--interval” argument. **Figure 1** shows a ranking using the *Sequence Prevalence Score* to prioritize variants from GISAID data processed in November of 2021, which is considered the initial month of emergence of the Omicron variant. In this ranking, the *Sequence Prevalence Score* for the Omicron variant is still relatively small but detectable.

**Figure 1.**
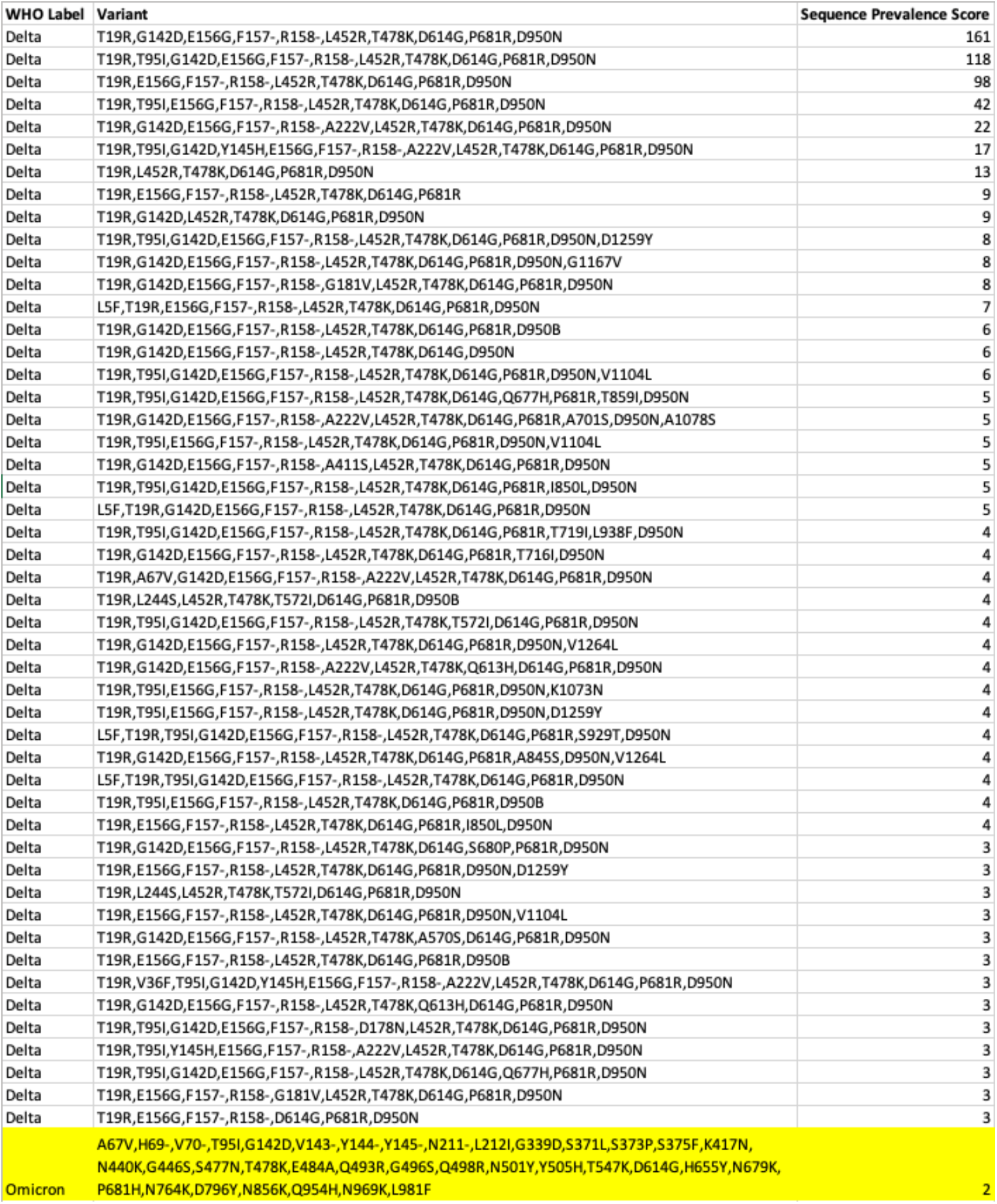
Global Spike Variant Ranking with the *Sequence Prevalence Score*. The output of a ranking based on GISAID data up to November 2021. The analysis returns a global ranking for all Spike protein variants ranked by the *Sequence Prevalence Score.* The initial emergence of the Omicron variant is being captured in this ranking, but at a relatively low rank, as highlighted in the table.

#### Functional Impact Score

While the Sequence Prevalence Score quantifies prevalence properties of variants, it requires some minimum time period to observe concerning trends and may not rank concerning variants high at early stages of emergence. A complementary approach for analyzing emerging variants independent of epidemiological dynamics is to predict their functional impact using prior knowledge from experimental datasets. We have designated regions of the Spike protein that have been experimentally shown to impact immune escape, receptor binding affinity, or viral replication as “Sequence Features of Concern” (SFoC). We detail our procedure for selecting SFoCs in the following section, but overall, these SFoCs include sites that could impact the binding of one or more monoclonal antibody classes^10^, neutralization with infection or vaccine (mrna-1273) induced antibodies, affect receptor binding affinity, or functionally important sites (NTD Supersite^13^, Furin Cleavage Site^28,29^, or D614G^16^).

Upon establishing our list of SFoCs, we assign each Spike variant a *Functional Impact Score* based on positional overlap with these critical regions. For each mutated position in the variant, if an overlap is found with one of the regions in the SFoC list, a score of 1 is assigned treating separately each of the monoclonal antibody classes, convalescent serum, vaccine serum, ACE2 binding affinity, NTD supersite, furin cleavage site, and site D614G. Any amino acid substitution or deletion that has a position falling within an SFoC is treated as one scorable mutation. Summing all the scores of each position mutated in the variant produces the *Functional Impact Score*. In **Figure 2** we provide a visual representation of what this positional overlap with SFoCs defined by deep mutational scanning antibody escape data would look like. The figure comes from an implementation of these annotations in the genome browser of the *BV-BRC SARS-CoV-2 Variant Tracker*^24^ and quantifies impact of monoclonal antibody binding by class for a mutation at each site on the Receptor Binding Domain.

**Figure 2.**
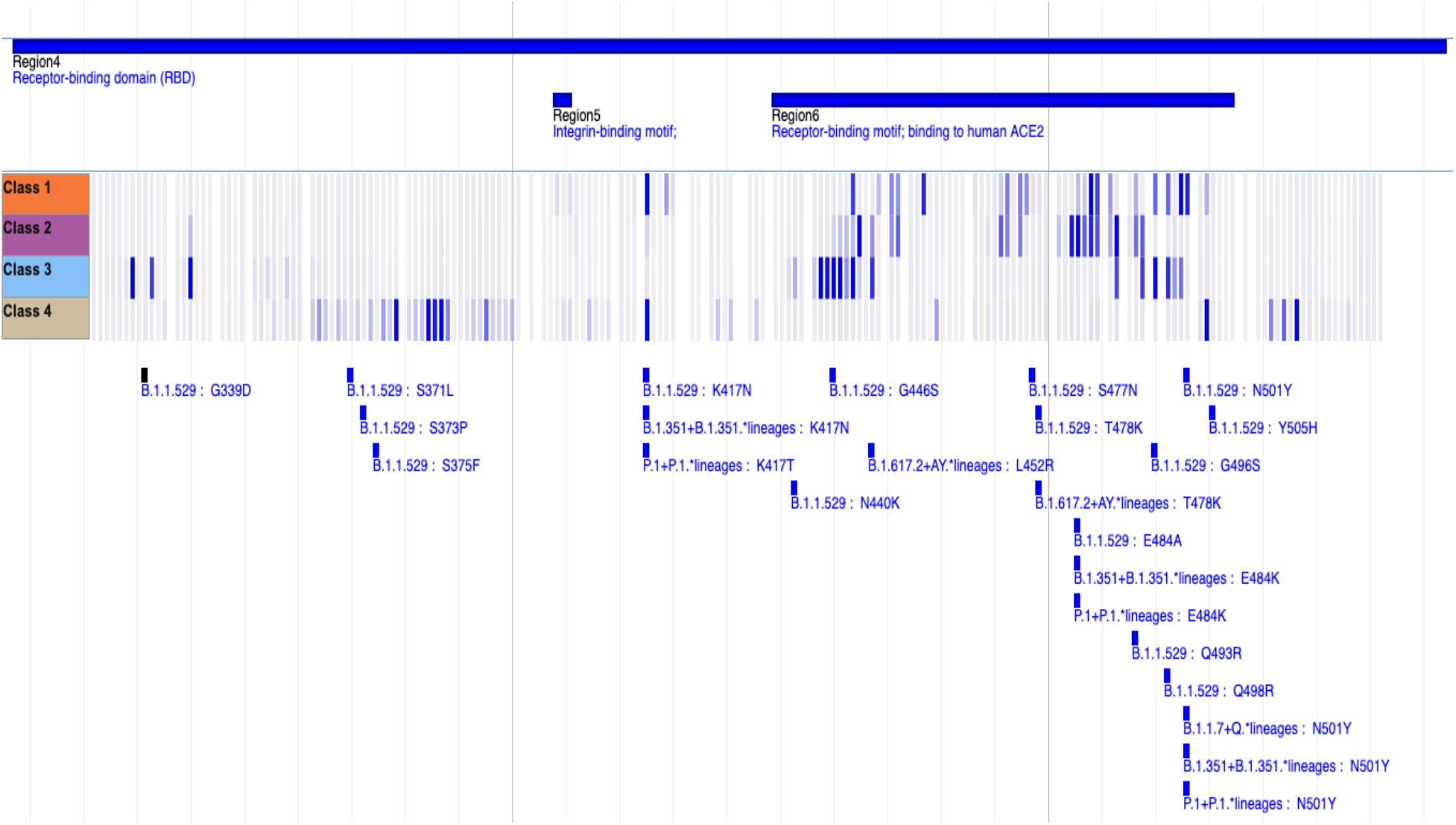
Sequence Features of Concern from Deep Mutational Scanning Antibody Escape Data. An image from the *BV-BRC SARS-CoV-2 Variant Tracker*^24^ genome browser shows a heatmap that quantifies the median escape fraction for each antibody class (Class 1 – 4), with the darker blue indicating greater escape fraction, implying greater antibody escape due to the mutation. We use these quantifications of potential antibody escape per mutated site per antibody class to define one type of Sequence Features of Concern. By identifying overlap between variants (bottom track) and these Sequence Features of Concern we can compute the *Functional Impact Score*.

#### Sequence Features of Concern

Most of the data used to define the SFoCs were derived from the deep mutational scanning experiments conducted by the Bloom Lab^2–9^. The resulting experimental data quantified the mutation impact towards monoclonal antibody escape, convalescent serum antibody escape, vaccine (mrna-1273) elicited antibody escape, and ACE2 binding affinity for nearly every position on the Receptor Binding Domain (RBD). To achieve these quantifications, the Bloom Lab constructed an RBD mutant library such that each amino acid site on the RBD was mutated with the 19 possible amino acid substitutions in the genetic background of the Wuhan-Hu-1 reference strain. The Bloom Lab used rigorous statistical processing to calculate an “escape fraction” for antibody escape (all between 0 and 1) and “binding average” for ACE2 affinity (all between −5 and 1) for every mutation at every position of the RBD. Each of the different monoclonal antibodies belonging to one of the four Barnes et al. structural epitope classes^10^, subject specific convalescent sera, and subject specific vaccine sera had their own escape fraction scores per mutation per site. The data from all these deep mutational scanning studies can be downloaded and explored at https://jbloomlab.github.io/SARS2_RBD_Ab_escape_maps/.

We examined the distribution of scores for each antibody escape datasets and the ACE2 binding dataset to narrow down on positions of the RBD vulnerable to significant increase in antibody escape or ACE2 binding affinity upon mutation. With a consolidated violin plot for all monoclonal antibodies and convalescent/vaccine sera, the analysis of antibody escape data showed that most of the escape fraction values were close to the 0 baseline and less than 0.2 (**Figure 3**). Therefore, to capture positions on the RBD that lead to strong antibody escape upon being mutated, an escape fraction threshold of 0.25 was applied. Consequentially, the antibody escape SFoCs were defined as RBD sites with one or more mutations that lead to an escape fraction exceeding this threshold for some monoclonal antibody, convalescent subject sera, or vaccine subject sera. The monoclonal antibodies corresponding to a mutation exceeding this escape fraction threshold were categorized into their structural epitope class to generalize the scoring for functional impact. As a result of this threshold, we designated 75 sites on the RBD that could significantly impact the binding of up to four antibody classes and 36 sites that could significantly impact the binding of antibodies from convalescent or vaccine sera.

**Figure 3.**
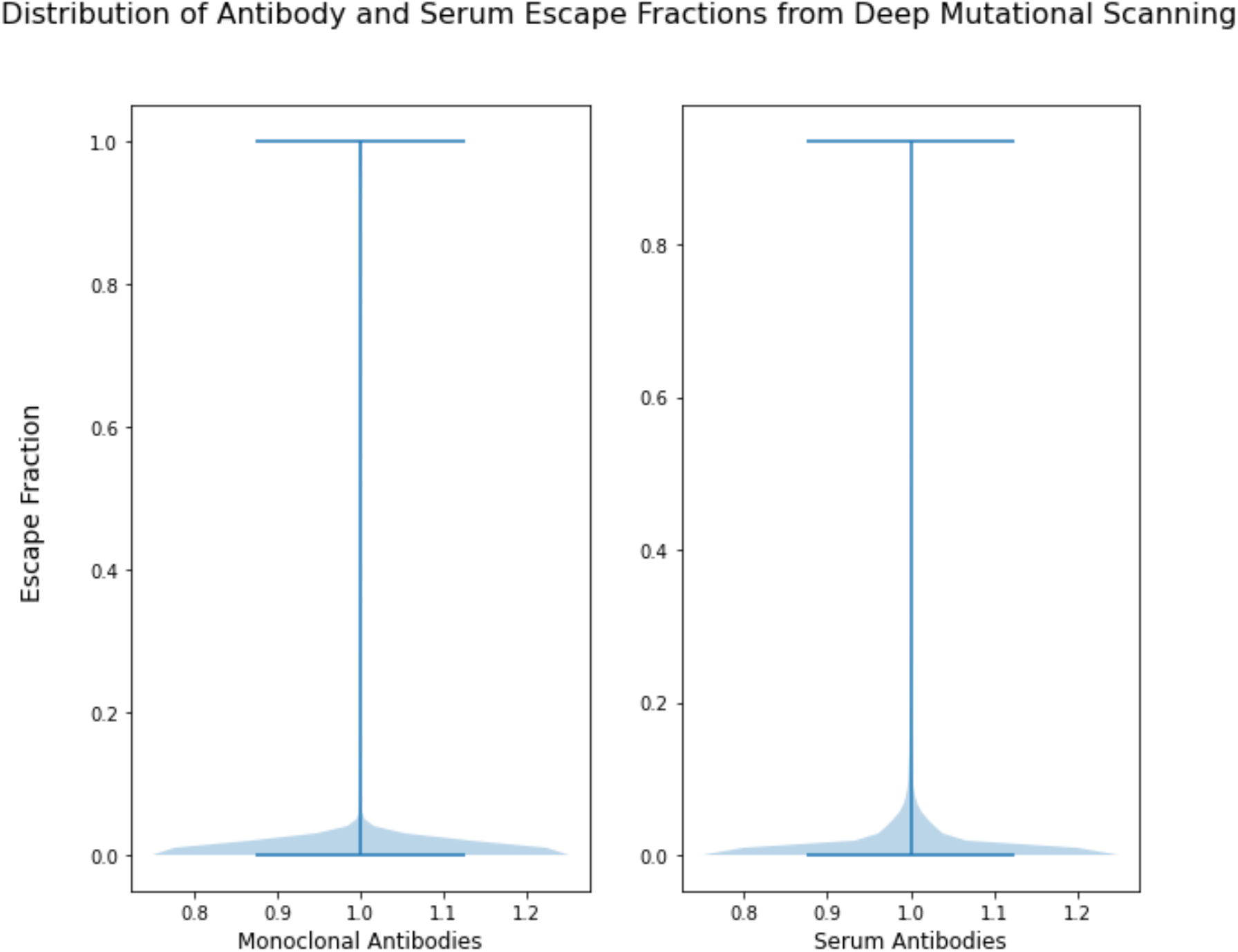
Defining Sequence Features of Concern from a Threshold of Escape Fractions. Violin plots of the distribution of escape fraction scores for monoclonal antibody escape and serum antibody escape from the Bloom Lab deep mutational scanning data. This distribution analysis led to an escape fraction threshold cutoff of 0.25 and subsequently led to the designation of 75 RBD sites significantly impacting monoclonal antibody binding and 36 sites impacting convalescent/Moderna vaccine (mrna-1273) sera elicited antibodies upon mutation.

Similarly, we evaluated the distribution of scores for the ACE2 binding affinity dataset. This analysis showed that most scores were close to the 0 baseline or negative, where a negative score implied a decrease in binding affinity (**Figure 4**). To identify mutations that led to increased ACE2 binding affinity, a binding average threshold of 0.1 was selected. As a result, each site with one or more mutations that exceeded this threshold was defined as an SFoC for increased ACE2 affinity, thus designating 12 sites that could significantly increase the binding to ACE2 upon mutation. We found site 501 to be included in this list, which is a notable site that leads to high degree of conformational alterations of the Spike RBD when bound to ACE2 upon acquiring a mutation^39^.

**Figure 4.**
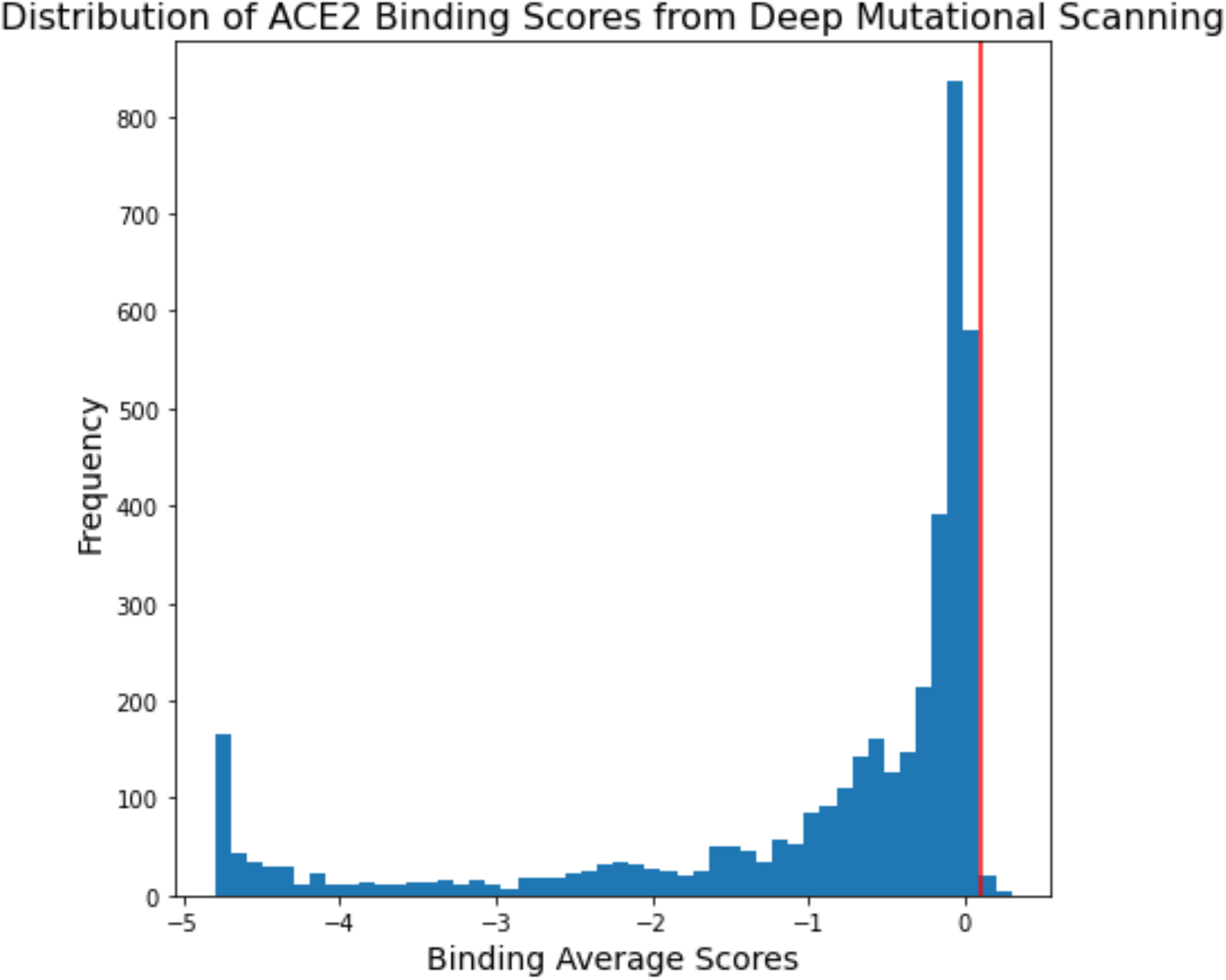
Defining Sequence Features of Concern from a Threshold of ACE2 Binding Scores. The distribution of ACE2 binding average scores from the Bloom Lab deep mutational scanning data. A score below 0 indicates a decrease in ACE2 binding affinity upon mutation, whereas a score above 0 indicates an increase in ACE2 affinity. Subsequently, this distribution analysis was used to select a threshold value of 0.1, that led to designating 12 RBD sites that could significantly increase ACE2 binding affinity upon mutation.

The remaining Sequence Features of Concern were those deemed critical for adaptive immunity or viral tropism, such as the NTD supersites^13^ (sites 14-20, 140-158, 245-264), site 614 of the Spike^16^, and the region flanking the furin cleavage site^28,29^ (sites 671-692). These were determined through literature curation.

#### Composite Score

Once the *Sequence Prevalence Score* and the *Functional Impact Score* have been computed, the two are summed to produce the *Composite Score*, which is used in our monthly reports to NIAID SAVE for variant rankings. *Composite Scoring* can be applied to all variants in a dataset, variants among a specified WHO clade, covariates among a specified PANGO lineage, or variants in a user supplied list. Thus, the *Composite Score* can simultaneously identify variants with alarming sequence prevalence dynamics ***and*** variants that would be predicted to impact important functional characteristics of the virus ***or*** both. For example, an initial analysis of the Omicron variant using epidemiological data from November 2021 did not show a high *Sequence Prevalence Score* (**Figure 1**), but the original Omicron sequence did show a very high *Functional Impact Score* and therefore a high *Composite Score* (**Figure 5**). These results highlight the importance of paying attention to both the *Sequence Prevalence Score* and *Functional Impact Score* for the early identification of Variants of Interest for further evaluation.

**Figure 5.**
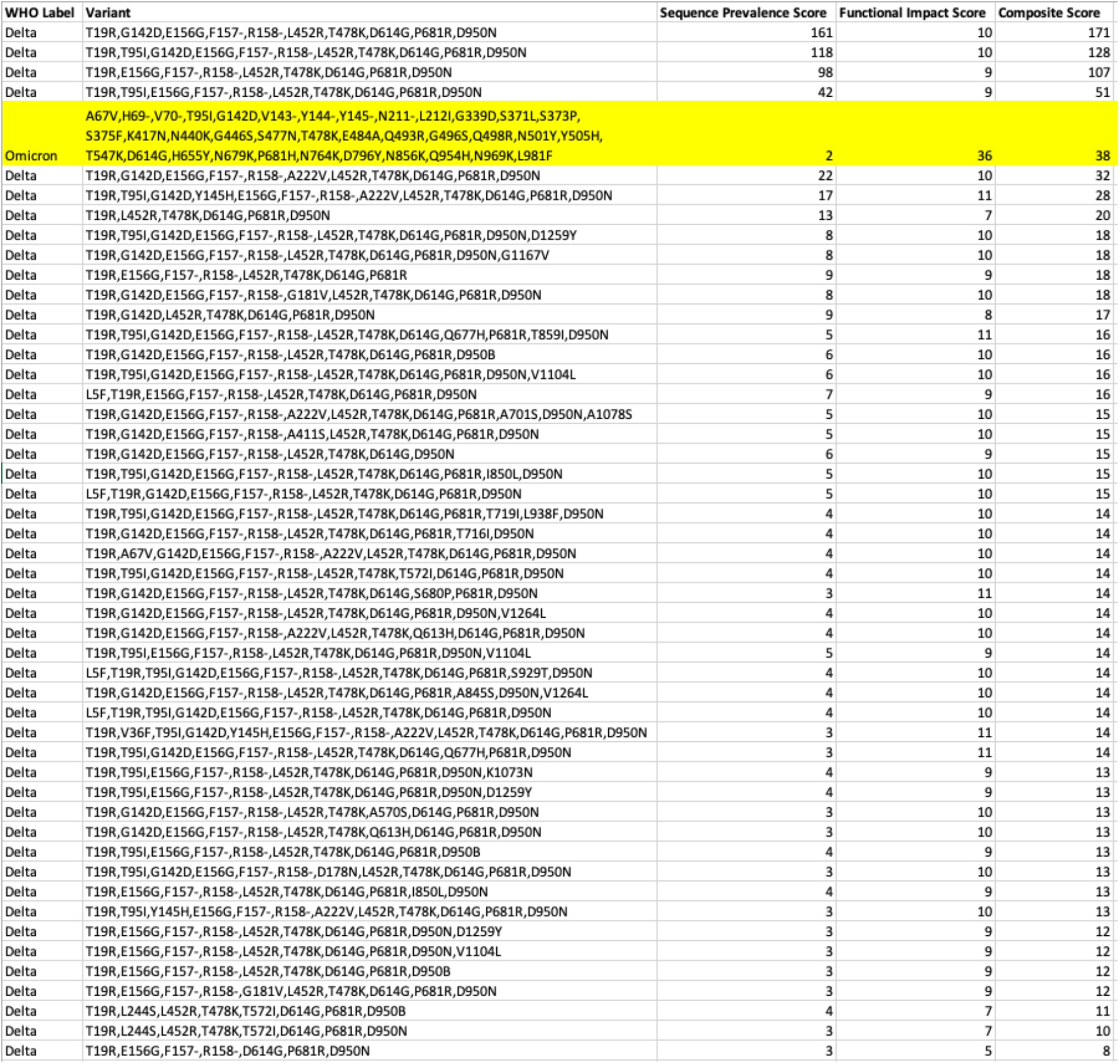
Global Spike Variant Ranking with the *Composite Score*. The output of a ranking based on GISAID data up to November 2021. The analysis returns a global ranking for all the Spike variants based on a *Composite Score*. In this case, the Omicron variant (highlighted in yellow) jumped considerably in rank relative to the *Sequence Prevalence Score* in Figure 1, thus showing the relevance of quantifying the *Functional Impact Score* in overall variant rankings.

The *Composite Score* is also very useful in quantifying subtle differences among covariates of the same clade. By January 2022, the most dominant lineage of Omicron clade was BA.1, which included several sub-lineages with varying Spike covariates, most significantly the addition of the R346K mutation. At the same time, BA.2 was also emerging and there was a primary variant constellation similar to BA.1 but with several unique mutation sites. When a list of these distinct Omicron covariates was supplied to the ranking algorithm, differences between the original BA.1 covariate and BA.1 + R346K could be observed through an increase in the functional impact score (**Figure 6**). In contrast, many of the other covariates had a *Sequence Prevalence Score* of 0, which indicated that those covariates were not displaying any significant regional growth and likely not as threatening as the original BA.1 or the BA.1 + R346K (now known as BA.1.1) covariates.

**Figure 6.**
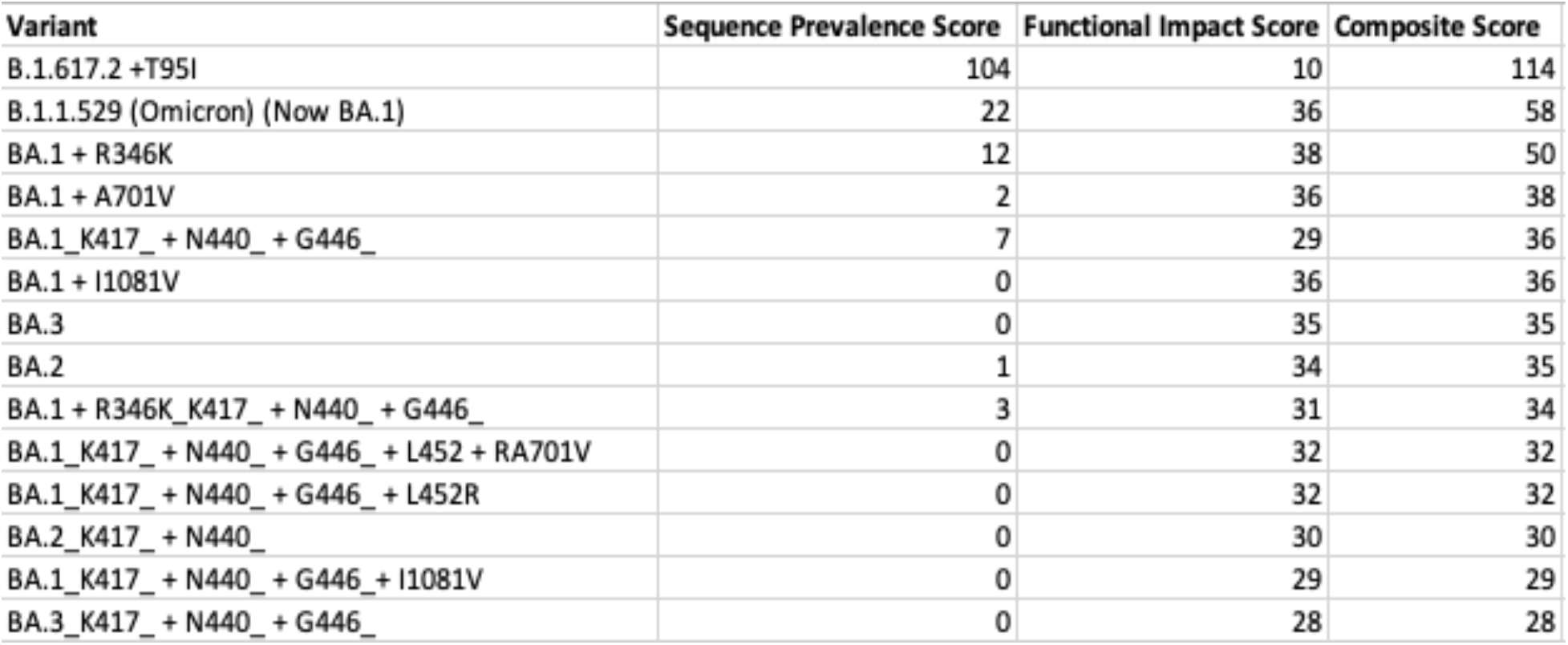
Global Spike Variant Ranking for a User Supplied Annotated List with the *Composite Score.* The output ranking from the *Composite Score* of an inputted list of covariates, annotated with names based on the additional or lost mutations relative to a consensus variant constellation, using GISAID data up to January 2022. These naming annotations offer simpler representations of the covariate sequence. The scoring in the last three columns quantitatively capture how these covariates differ, including the increase in functional impact for BA.1 + R346K relative to the ancestral BA.1.

#### Mutation Prevalence Score

In addition to the covariate constellation analysis, a *Mutation Prevalence Score* analyzing single amino acid substitutions on the SARS-CoV-2 Spike protein is also calculated. As with the *Sequence Prevalence Score*, this approach uses data from the past three months to assign a score of 1 for every country/month combination in which the prevalence of an amino acid mutation is >5% or the growth rate is greater than 5-fold. **Figure 7A-B** show the results of ranking mutations within the RBD and NTD using global GISAID sequence data from December 2021.

**Figure 7.**
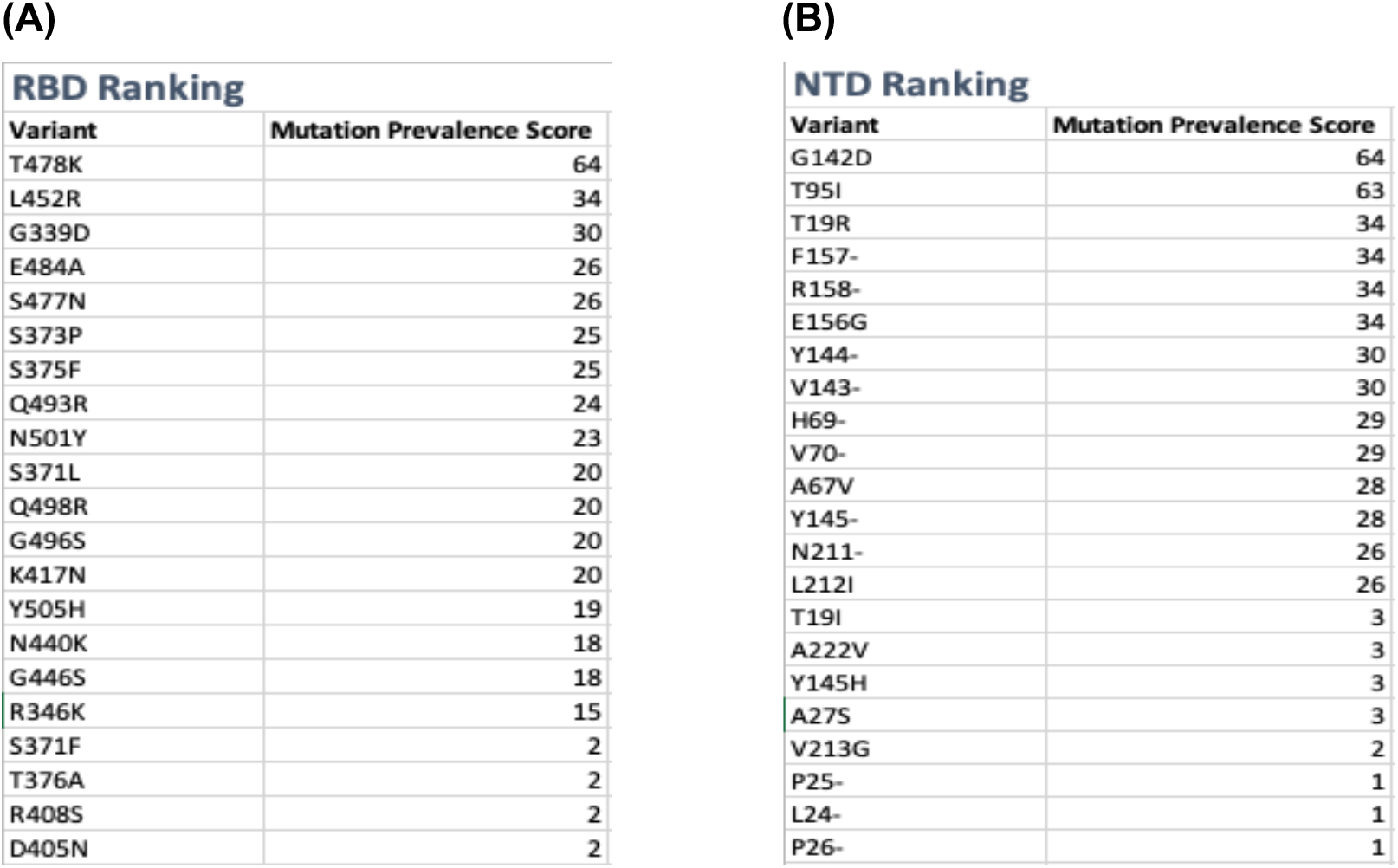
Global Single Amino Acid Mutation Ranking with the *Mutation Prevalence Score*. **(A)** The output of an RBD mutation ranking and **(B)** the output of an NTD mutation ranking based on GISAID data up to December 2021. The results return a ranking for Spike protein amino acid mutations ranked by the *Mutation Prevalence Score* within their respective domain, RBD or NTD.

#### Emerging Lineage Score

In some cases, researchers may just want to know how the various PANGO Lineage designations differ in terms of their epidemiological dynamics. As we saw with Delta, there were a multitude of AY.* lineages for the entire WHO clade designated in a relatively short time frame, so ranking these lineages would be helpful to identify which should be prioritized for further analysis.

As with the *Sequence Prevalence Score*, the *Emerging Lineage Score* begins by filtering for covariates with an assigned country and with a variant count greater than 10 in the most recent month. From there, using the past three months of data, for each PANGO lineage, every unique covariant/country/month combination in which the growth rate is greater than 15 is assigned a score of 1; these values are summed to compute the *Emerging Lineage Score*. Since this algorithm is counting multiple distinct covariates comprising each lineage, it could be biased towards PANGO lineages with very abundant covariates; hence, we used a higher growth rate threshold to capture the key covariates driving the overall growth of the lineage. A growth rate of 15 was chosen as it results in a relatively stable list of PANGO lineages. Growth rates above 15 did not significantly affect the ranking of the list. This growth rate is also appealing in that it is relatively high and therefore relatively stringent. **Figure 8A-B** shows the results from this method using GISAID data from December 2021 and January 2022 to rank lineages globally. BA.1 was the dominant Omicron lineage in December 2021 and was the lineage with the highest *Emerging Lineage Score*. (**Figure 8A**). However, by January 2022 additional Omicron lineages were rapidly growing with the presence of BA.1.1 and BA.2 (**Figure 8B**). Since these lineages are made up of multiple covariates, we could take the results returned from the *Emerging Lineage Score* to decide which covariates within these lineages warranted further investigation by running a PANGO lineage specific *Composite Score* ranking to prioritize the covariates within a specific lineage, as shown in **Figure 9** with BA.2, which captured a single covariate of the lineage with the strongest dynamics as early as January 2022.

**Figure 8.**
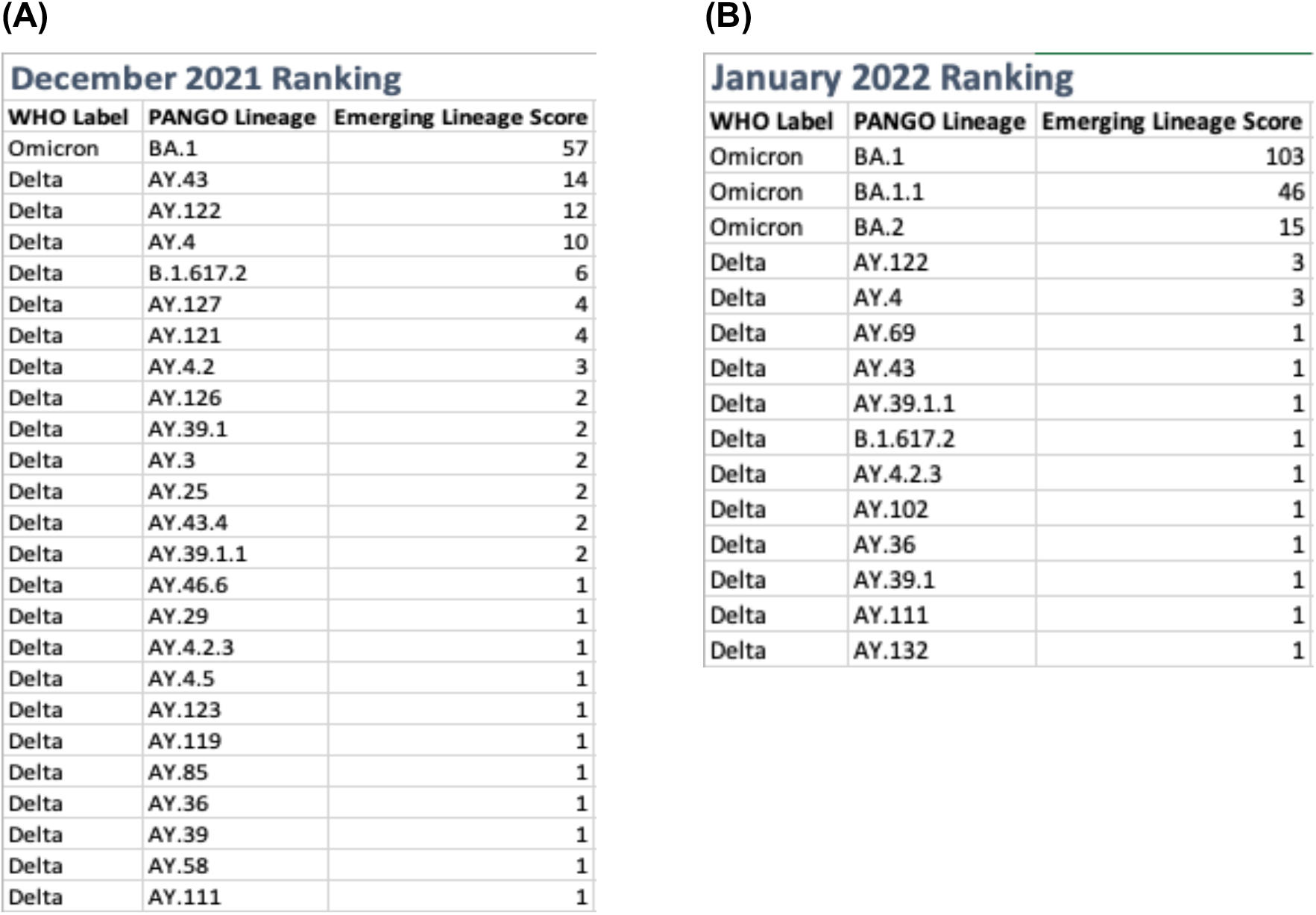
PANGO Lineage Ranking with the *Emerging Lineage Score.* The output of a lineage ranking based on GISAID data up to December 2021 **(A)** and January 2022 **(B)**. The results return a global ranking of PANGO Lineages based on the *Emerging Lineage Score.* In December 2021, BA.1 was the strongest emerging lineage with other Delta sub-lineages still on the rise. However, by January 2022, several Omicron lineages were newly emerging, with the Delta lineages tapering off.

**Figure 9.**
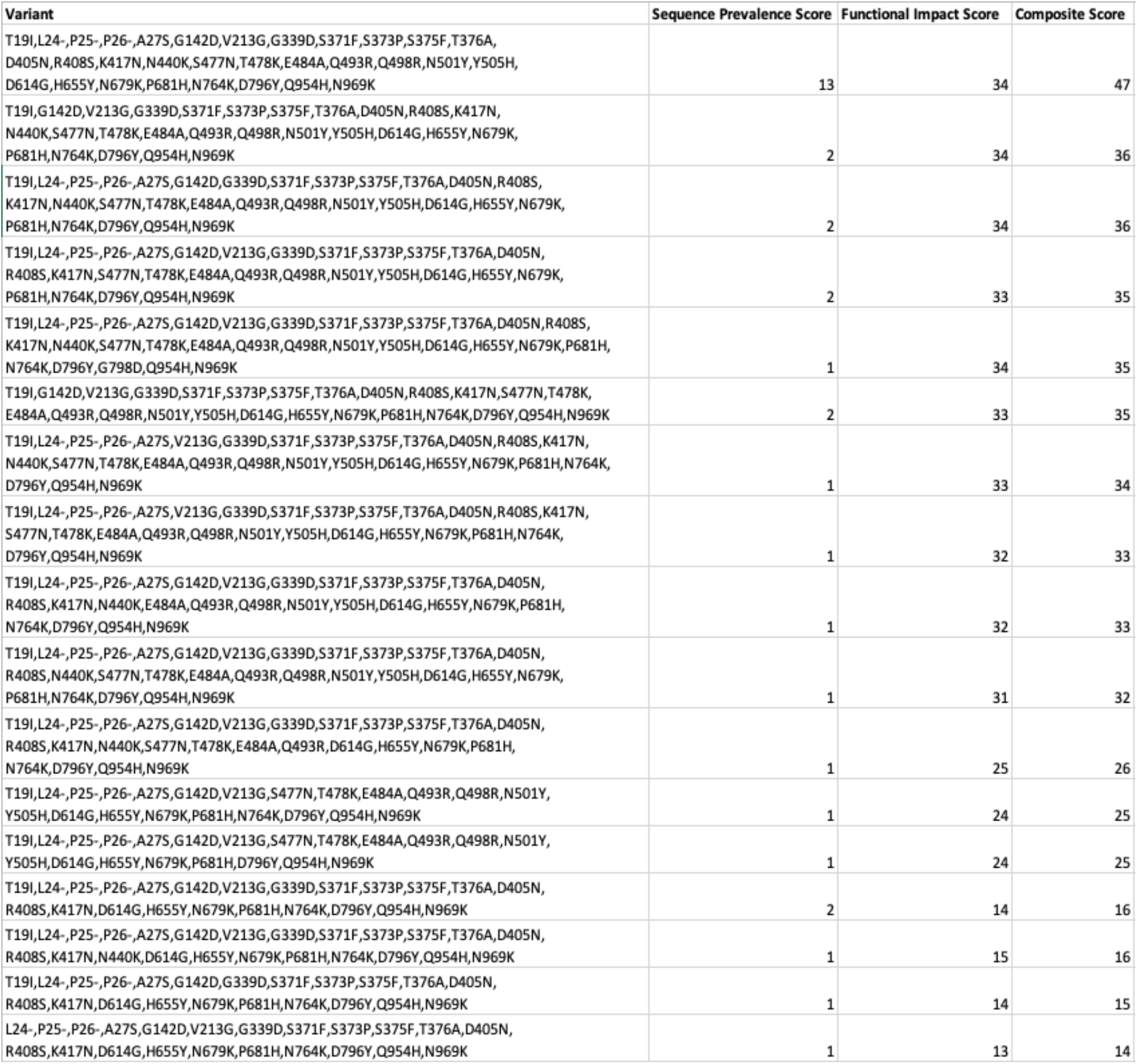
Global Ranking of BA.2 Variants with the *Composite Score*. The output of a *Composite Score* ranking based on GISAID data up to January 2022. The purpose of this ranking is to focus the analysis on covariates within a specific lineage, in this case within BA.2. The ability to capture a dominant covariate (top row) likely driving much of the observed dynamics for this lineage can be observed.

### Visualizing Early Detection

To complement our early detection analysis of emerging SARS-CoV-2 variants through our scoring and ranking algorithms, we found it useful to visualize variant growth both globally and regionally to further understand the dynamics of these variants and facilitate early detection. We demonstrate the utility of these visualizations for early detection purposes by providing some case studies regarding the early dynamics of the Delta and Omicron variants.

#### Visualizing Relative Growth of PANGO Lineages

When a new variant displays alarming epidemiological dynamics or predicted functional changes, like Delta and Omicron did, researchers may want to visualize the growth dynamics of the new variant in the context of how the prevalence of other variants are changing. We commonly see during viral evolution that when a new dominant variant/strain emerges, it triggers a phenomenon where the growth of the currently circulating variants suddenly begin to sharply decline, perhaps because the new variant has a fitness advantage and is able to outcompete the older variants^16,19,31,33^. If we observe that the prevalence of a novel variant with alarming characteristics is increasing with a corresponding sudden decline in growth of other circulating variants, this will further indicate the early detection of a potential Variant of Concern. As an example, prior to the emergence of Delta, B.1.1.7-derived Alpha variants were the most dominantly circulating variants around the world. However, between May and June of 2021, it was becoming clear that the newly emerging Delta variant was displaying noteworthy properties^19^ and very quickly replacing the other circulating variants, including B.1.1.7 (Alpha), both globally (**Figure 10A**) and regionally, especially in the UK (**Figure 10B**). By visualizing these dynamics in stacked line plots, the relative magnitude by which a novel variant is growing offers further evidence of the early detection of a potential Variant of Concern.

**Figure 10.**
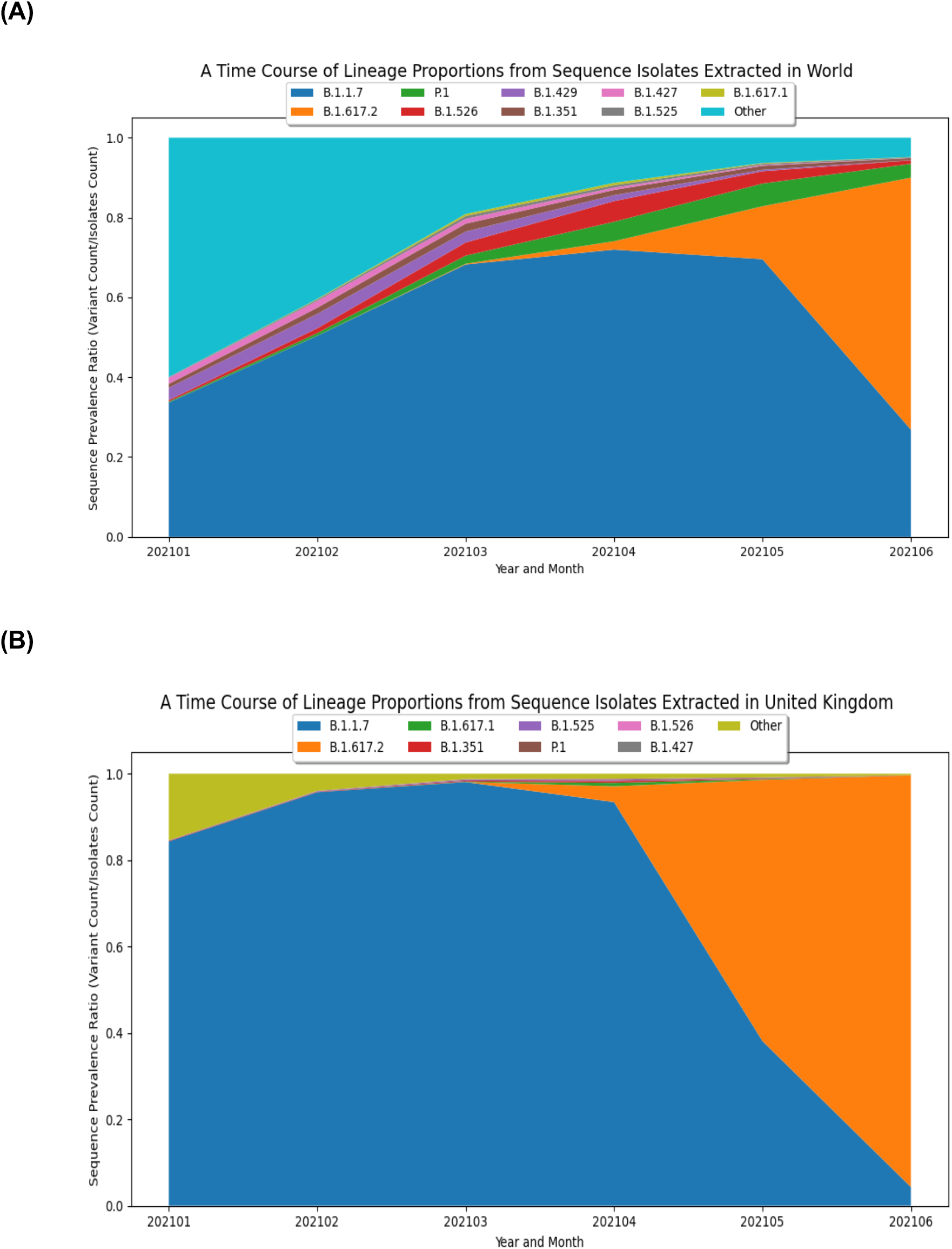
Visualizing the Emergence of the Delta (B.1.617.2) Variant. **(A)** A plot of global trends overtime based on GISAID data up to June 2021 to visualize the early growth dynamics of Delta. The graph displays the 10 PANGO lineages with the most substantial global prevalence dynamics over a six-month time frame, with the early emergence of B.1.671.2 and the sudden sharp decline of B.1.1.7 clearly evident. **(B)** A plot of growth trends for Delta in the United Kingdom based on GISAID data up to June 2021. The graph displays the PANGO lineages with the most substantial global prevalence dynamics over a six-month time frame solely within the United Kingdom and shows the local growth of B.1.617.2 and decline of B.1.1.7.

#### Visualizing Growth of Variants

In addition to tracking the emergence and growth of new PANGO lineages, it is also useful to visualize the evolution and growth of variants within the lineage. As discussed earlier, B.1.1.529 (Omicron) started to show rapid growth in December 2021 and then quickly accumulated additional substitutions, ultimately generating the newly designated BA.1 and BA.2 lineages and sub-lineages. By providing a list of covariates of interest, like the one used in **Figure 6**, our pipelines produced a graph showing the prevalence of a selected list (**Figure 11**). Comparing the *Composite Score* results from **Figure 6** with the visualization in **Figure 11**, some interesting insights about BA.1 + R346K emerge. BA.1 + R346K had a high functional impact score, and while the *Sequence Prevalence Score* was also high, it wasn’t quite as high as BA.1 for the month of January 2022. However, the **Figure 11** visualization shows that BA.1 + R346K was exhibiting a sharper change in its global prevalence relative to BA.1, suggesting that this variant would warrant further monitoring. Indeed, these plots provide a complementary representation to the *Composite Score* ranking to facilitate early detection analysis and more confidently identify variants that warrant experimental evaluation.

**Figure 11.**
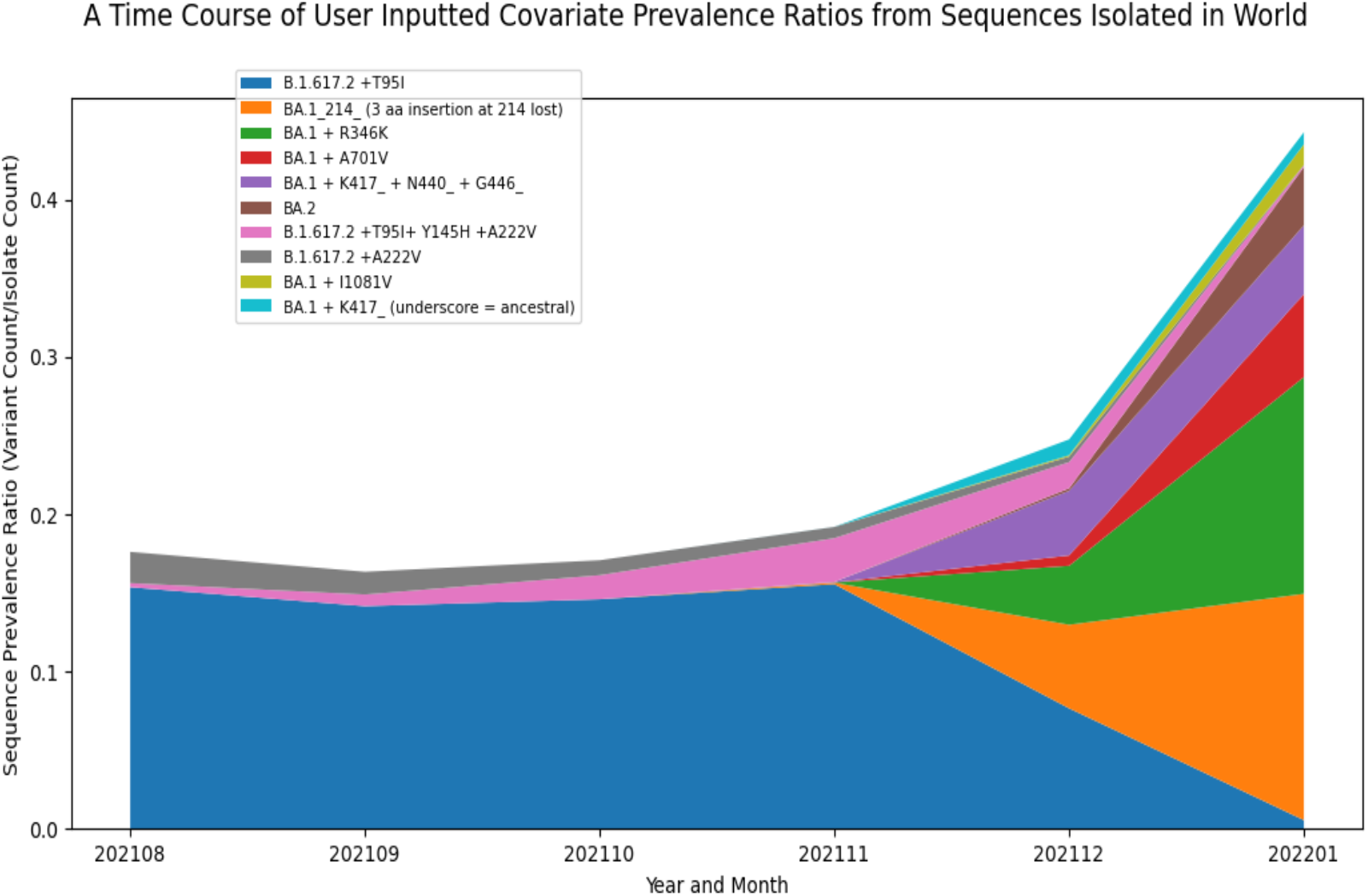
Visualizing the Emergence of Omicron Variant Constellations. A plot displaying the prevalence dynamics over six months of Spike covariates in an inputted file based on GISAID data up to January 2022. This visualization captures the sharp growth and relatively large global prevalence of BA.1 + R346K. The names presented in the legend mean the ancestral PANGO lineage +/- certain amino acid mutations to represent different covariates. Underscores indicate that certain mutations were lost relative to the ancestral sequence.

#### Visualizing Growth of Individual Amino Acid Substitutions

Analysis of the characteristics of lineages and covariates also provided some insights into the different amino acid mutations that contribute to the multitude of covariates, in some cases from different lineages, perhaps due to convergent evolution or recombination. To assess their individual effects, the dynamics of selected single amino acid substitutions can also be plotted. For example, prior to January 2022, the L24-, R346K, N440K, G446S, L452R, A701V mutations were appearing sporadically throughout our ranked covariates. Plotting the dynamics of these individual amino acid mutations over subsequent time periods shows that the R346K and G446S mutations started to decrease in prevalence at the same time as the prevalence of L24- was rapidly increasing, suggesting that viruses carrying this mutation may possess a fitness advantage (**Figure 12**). Indeed, this L24- is part of an extended deletion that distinguishes BA.2, which subsequently replaced BA.1.

**Figure 12.**
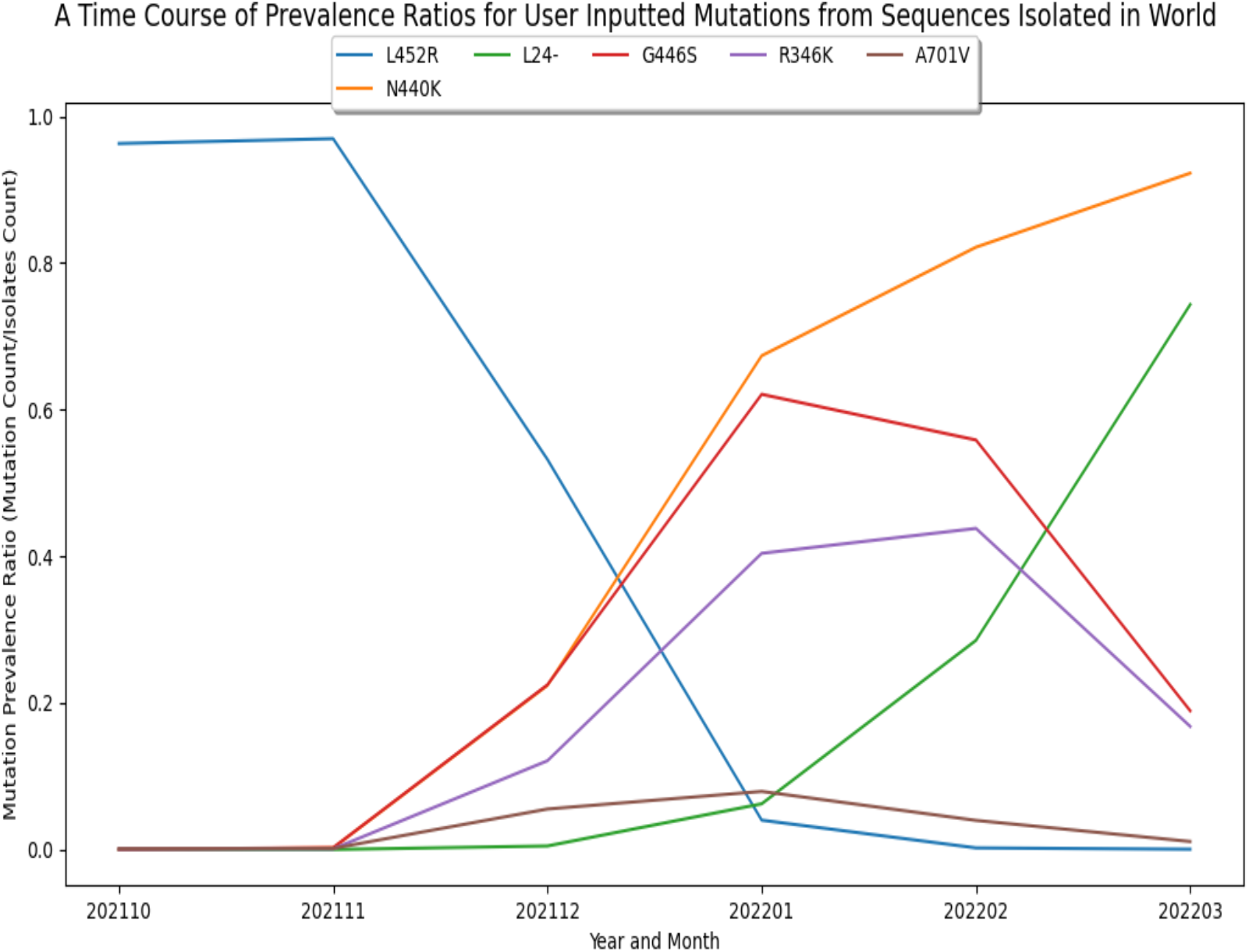
Visualizing Growth Patterns of Single Amino Acid Mutations in the Spike Protein. A plot based on GISAID data up to March 2022 demonstrating the shift in dynamics for individual amino acid mutations. This plot is based on an inputted list of six amino acid mutations. Note that the L24- is a part of an extended deletion that also includes P25 and P26 found in BA.2.

### Challenges

Defining SARS-CoV-2 variants that warrant experimental evaluation for functional testing presents significant challenges. Early genomic sequencing data is often subject to data imbalances or biases with respect to specific geographic regions. Wealthier countries with higher sequencing capacity, such as the US and UK, are responsible for the majority of publicly deposited SARS-CoV-2 genomes. Of the 4.5 million SARS-CoV-2 genomes available in GenBank and BV-BRC as of April 2022, roughly 47% were from the US and 38% from the UK. Another challenge is the sequence ambiguity that can occur as a new variant emerges and before the sequencing assays can be optimized. And since the SARS-CoV-2 genomes submitted to the public databases have already been assembled, the quality of the read level data cannot be easily evaluated independently. Processing variant data through the GISAID metadata file downloads, many sequences appeared to have reverted to ancestral residues in comparison to the original Omicron outbreak sequences, but in many cases this is due to low sequence coverage in certain genomic regions that is not apparent in the metadata file. Indeed, processing assembled GenBank sequencing data from BV-BRC, only about 25% of sequences had little to no ambiguities, and the amount of ambiguity in the sequencing data fluctuated during the initial emergence of certain important variants, like Delta and Omicron, making it challenging to compute true sequence prevalence of authentic covariates. Another challenge is choosing the amount of data that ought to be regularly downloaded for computing the early detection scoring heuristics. While focusing on the most recent data could potentially be used to identify concerning variants more swiftly, a potential drawback would be sequence biases resulting in minimal representation. On the other hand, longer temporal data is more comprehensive and accurate, but could delay identification of newly emerging variants of concern. In our pipeline, we allow the option to choose anywhere from the past 2 – 6 months of global sequencing data to evaluate, with a default of 3 as our best attempt to set a balance between early detection and unbiased, accurate results. Finally, the most enduring challenge is the fact that these data are very large and continuously growing, as new SARS-CoV-2 sequencing data are being deposited by the thousands every day. Designing pipelines to carryout real-time genomics analysis for a shear amount of data is a technically challenging task, and our techniques on how best to manage, analyze, and scale will need to continuously adapt.

## Discussion

Over two and a half years since the initial declaration of the COVID-19 pandemic, it seems clear that SARS-CoV-2 will persist in our community and remain a public health issue for the indefinite future. However, if we, as a biomedical community, come together and maximize our resources to combat this virus, we can consistently minimize the threat it brings to our world and transition to a phase in which SARS-CoV-2 becomes an endemic infection with only modest effects on public health. A major factor that could contribute to consistently minimizing the threat brought by SARS-CoV-2, or any other emerging pathogen, is through genomic surveillance. Our most precise approach to monitoring viral evolution to ensure that we maintain reliable therapeutics and accurate diagnostics is by routinely collecting samples from infected individuals and using sequencing technologies to acquire the whole genome sequences. With the COVID-19 outbreak, the research and public health communities have truly excelled at this task, as we have now reached a point where millions of SARS-CoV-2 genome sequences have been deposited in public databases. That being said, all of these data are only as powerful as the computational resources used to analyze them. Thus, this explosion in publicly available viral genomes also calls for the development of appropriate computational frameworks that can process and scale as the data grows to enable the timely identification and prioritization of emerging variants.

In this work, we present approaches to process SARS-CoV-2 genomic sequences along with epidemiological metadata on a regular basis and apply scoring heuristics to prioritize variants based on their epidemiological dynamics and predicted functional characteristics by computing *Sequence Prevalence, Functional Impact,* and the *Composite Scores*. The output provides concise lists of ranked variant constellations (covariates), offering a straightforward approach for wet-lab scientists to immediately determine which combinations of mutations ought to be evaluated experimentally. These methods were validated through the detection of the Omicron variant and provided a relatively high-ranking score at early stages of emergence in November 2021. In addition to early detection of novel variants like Omicron, our system makes it easy to evaluate the subtle differences among the multitude of covariates arising from a single lineage, like comparing the BA.1 + R346K covariate to the original BA.1 covariate. Finally, we also provide multiple visualization methods for variant growth either by PANGO lineage, covariate, or single amino acid substitution and demonstrate how coupling rankings with visualizations can further ground our confidence in early detection of variants that warrant experimental evaluation.

While the initial focus of this work was on Spike protein variants, as that was NIAID SAVE’s interest for evaluation, the framework was recently extended to score and rank proteome-wide SARS-CoV-2 variant constellations and single amino acid substitutions, which we provide in our publicly available pipeline. While the primary focus was the Spike protein due to its critical role in adaptive immunity and viral tropism, the community is beginning to take serious interest in mutations arising in non-structural proteins, particularly nsp3, nsp5, and RNA-dependent RNA Polymerase (nsp12), due to their importance in the SARS-CoV-2 replication cycle and antiviral drug targeting^34–38^. We are paying close attention to the literature to monitor the science behind non-Spike protein regions that play key roles in replication or impact drug targeting, and are continually updating our Sequence Features of Concern to account for this new knowledge. Ultimately, our framework can begin providing *Composite Scores* for a variant constellation specific to any SARS-CoV-2 protein.

The methods presented in this work could be extended for evaluating variants of other viral species if sufficient data granularity is available. This approach requires enough genomic sequencing data, consistent spatiotemporal isolation metadata, a methodology to compute variants with respect to a reference or a consensus genome^22^, and sufficient prior knowledge through experimental data to define Sequence Features of Concern and quantify functional impacts. Moreover, these algorithms could be extended to evaluate genomic surveillance data from zoonotic disease reservoirs, such as influenza virus in avian or swine species. Indeed, several projects already exist to collect viral genomic sequences from such reservoirs and warehouse these data in public databases^18^. Overall, the methodologies described here can play an important role in a complete public health ecosystem by utilizing genomic sequencing data to monitor viral evolution and remain steps ahead of SARS-CoV-2, or any other virus, and ultimately deter the next pandemic.

## Data Availability

Publicly available datasets were analyzed in this work, and the original contributions presented are included in the manuscript. Further inquiries for data availability can be directed to the authors.

https://github.com/zwallace425/SARS-CoV-2_Pipeline

## Data Availability Statement

Publicly available datasets were analyzed in this work, and the original contributions presented are included in the article/**Supplementary Material**. Further inquiries can be directed to the corresponding author.

## Author Contributions

ZSW conceptualized the methods, collected/analyzed data, developed the public version of the pipeline, and wrote the original draft of the manuscript. JD, RDO, RS, and MS conceptualized the methods and contributed to pipeline development. AN, YZ and CMZ conceptualized the methods and reviewed and edited the manuscript. RHS was the principal investigator who conceived the project, conceptualized the methods, supervised the project, acquired funding, and reviewed and edited the manuscript.

## Acknowledgements

We acknowledge the Bloom Lab at Fred Hutchinson Cancer Research Center for their advice and data sharing, the Korber Lab at Los Alamos National Laboratory for their advice on methodology, Fritz Orbermeyer/Lemieux Lab at the Broad Institute for code sharing, the San Diego Supercomputer Center/XSEDE for the allocation of the Expanse compute resources, and both the originating and submitting laboratories for the sequence data deposited in INSDC and GISAID databases. The project has benefited from the ongoing discussion among the computational biologists and experimentalists in the NIAID SAVE consortium and among the analysts and developers of the BV-BRC.

## Supplementary Material

The *BV-BRC SARS-CoV-2 Early Detection and Analysis Pipeline*, the list of Sequence Features of Concern, and a more detailed version of the algorithms along with pseudocode are available from https://github.com/zwallace425/SARS-CoV-2_Pipeline.

